# Association of a pace of aging epigenetic clock with rate of cognitive decline in the Framingham Heart Study Offspring Cohort

**DOI:** 10.1101/2024.05.21.24307683

**Authors:** M.J. Savin, H. Wang, H. Pei, A.E. Aiello, S. Assuras, A. Caspi, T.E. Moffitt, P.A. Muenning, C.P. Ryan, B. Shi, Y. Stern, K. Sugden, L. Valeri, D.W. Belsky

## Abstract

**Introduction:** The geroscience hypothesis proposes systemic biological aging is a root cause of cognitive decline.

**Methods:** We analyzed Framingham Heart Study Offspring Cohort data (n=2,296; 46% male; baseline age *M*=62, SD=9, range=25-101y). We measured cognitive decline across two decades of neuropsychological-testing follow-up. We measured pace of aging using the DunedinPACE epigenetic clock. Analysis tested if participants with faster DunedinPACE values experienced more rapid preclinical cognitive decline as compared to those with slower DunedinPACE values.

**Results:** Participants with faster DunedinPACE had poorer cognitive functioning at baseline and experienced more rapid cognitive decline over follow-up. Results were robust to confounders and consistent across population strata. Findings were similar for the PhenoAge and GrimAge epigenetic clocks.

**Discussion:** Faster pace of aging is a risk factor for preclinical cognitive decline. Metrics of biological aging may inform risk stratification in clinical trials and prognosis in patient care.

## 1. INTRODUCTION

As we grow older, we experience a progressive loss of integrity and resilience capacity in our cells, tissues, and organs^1^. Within the emerging field of Geroscience, this process is referred to as “biological aging,” and is thought to originate from an accumulation of molecular damage^2^ that manifest as a series of cellular-level changes or “hallmarks of aging.”^3^ These hallmarks of aging in turn are implicated in the etiology of many different chronic diseases, including Alzheimer’s disease and related dementias (ADRD)^4,5^.

One of the strongest phenotypic risk factors for ADRD is preclinical cognitive decline, a more rapid decline in cognitive abilities prior to meeting diagnostic criteria for mild cognitive impairment^6^. In observational cohort studies, the rate of preclinical cognitive decline varies, with some individuals maintaining healthy cognitive function for many years while others decline rapidly to ADRD onset^7,8^. Better understanding of the causes of this variation are needed to inform risk stratification in clinical trials and improve prognosis in clinical care^9^. Here we test the geroscience-informed hypothesis that some individuals experience more rapid preclinical cognitive decline than others because they have a faster pace of biological aging.

We analyzed data from the Framingham Heart Study Offspring Cohort. We modeled trajectories of cognitive decline from two decades of neuropsychological testing data. We measured pace of biological aging from DNA methylation data using the DunedinPACE epigenetic clock. We tested if participants with faster pace of aging exhibited accelerated cognitive decline. We evaluated robustness of results across specifications considering a range of confounders and effect modifiers, including smoking history, cell composition of blood samples used to derive DNA, presence of Mild Cognitive Impairment (MCI) at baseline, level of cognitive functioning at baseline, sex, and APOE4 carrier status. Finally, we tested if pace of aging associations with cognitive decline reflected a process contributing to risk of dementia. We repeated analysis for two other proposed metrics of biological aging, the PhenoAge and GrimAge epigenetic clocks.

## 2. METHOD

### 2.1 Participants

The Framingham Heart Study (FHS) is an ongoing population-based cohort following three generations of families recruited, starting 1948, within the town of Framingham, Massachusetts, USA. We analyzed data from the second generation of participants, the Offspring Cohort. The Offspring Cohort (*N*=7306) was initiated in 1971 and participants have since been followed-up at nine examinations, approximately every 4-7 years. The study protocol was approved by the institutional Review Board for Human Research at Boston University Medical Center, and all participants provided written inform consent. Data for the Framingham Offspring Study were obtained from dbGaP (phs000007.v33.p14).

Our analysis focused on measurements of biological aging from DNA methylation (DNAm) data collected at the 8^th^ follow-up visit and measurements of cognitive decline from neuropsychological test data collected beginning around the time of the 7^th^ study visit and ongoing through 10 years after the 9^th^ study visit **(Supplementary** Figure 1).

### 2.2 Biological Aging

While the hallmarks of aging themselves are difficult to measure in human observational studies, methods based on machine-learning have spawned a range of new biomarkers of biological aging from analysis of -omics data^10^. The best-validated of these omics biomarkers are a family of DNAm algorithms known as epigenetic clocks^11^. The newest generation of epigenetic clocks show strong associations with aging-related morbidity/mortality and appear to capture the wear and tear arising from environmental and social determinants of health.^12,13^. Among this new generation of epigenetic clocks, the most consistent predictor of cognitive functioning and risk for dementia is DunedinPACE. Within the Dunedin Study cohort, those with more rapid decline in the longitudinal-change phenotype exhibited more rapid cognitive decline and signs of accelerated brain aging in midlife^14,14–16^. Beyond the Dunedin Study, children and adults with faster pace of aging as measured by the DunedinPACE epigenetic clock tend to perform more poorly on cognitive tests as compared to age-peers with slower pace of aging^17–19^ and to exhibit signs of accelerated brain aging^20,21^. Given the basis of this prior evidence, we focus analyses on DunedinPACE as our primary independent variable. We include, for comparison, analysis of the two other epigenetic clocks with robust evidence for prediction of morbidity and mortality, PhenoAge^22^ and GrimAge^23^, although prior evidence of their association with ADRD is inconsistent^24^.

DNA methylation was measured from whole-blood DNA samples collected at the eighth study visit. Assays were performed with the Illumina 450K Array at the University of Minnesota and John Hopkins University (dbGaP phs000724.v9.p13). Array data from both sites were pooled and processed from raw IDAT files by the Geroscience Computational Core of the Robert N. Butler Columbia Aging Center. After quality control, data were available for *n*=2296 participants. Details of the preprocessing and quality control steps employed are reported in the **Supplemental Methods.**

#### 2.2a DunedinPACE

The DunedinPACE epigenetic clock is a measure of the pace of biological aging^25^. It was developed from analysis of longitudinal change in 19 biomarkers of the integrity of the cardiovascular, metabolic, renal, hepatic, immune, dental, and pulmonary systems over a 20-year follow-up period in the Dunedin Study birth cohort. Initiated in 1972-3, the Dunedin Study followed a single-year birth cohort across five decades. Biomarker measurement was conducted when participants were 26, 32, 38, and 45 years of age. The DunedinPACE algorithm was developed by first modeling change over the 20 years of follow-up to create a composite Pace of Aging phenotype^14,15,16^. Next, Pace of Aging was modeled from whole-blood DNA methylation measured at the age-45 follow-up to derive the DunedinPACE algorithm^25^.

DunedinPACE has an expected value of 1 in midlife adults, corresponding to a rate of 1 year of biological aging per 12-months of calendar time. Values >1 indicate a faster pace of aging (e.g. a value of 1.25 would indicate a pace of aging 25% faster than the norm for midlife adults); values <1 indicate a slower pace of aging (e.g. a value of 0.75 would indicate a pace of aging 25% slower than the norm for midlife adults). We computed DunedinPACE in Framingham Heart Study participants using the R package available from GitHub (https://github.com/danbelsky/DunedinPACE). For analysis, DunedinPACE values were scaled to have *M*=0 and *SD*=1.

#### 2.2b Other Epigenetic Clocks

Other candidate measures of aging can be computed from DNA methylation data. For comparison, we repeated analysis with the PhenoAge and GrimAge epigenetic clocks. In contrast to DunedinPACE, which measures pace of aging, GrimAge and PhenoAge are static measures of biological age^22,23^; whereas DunedinPACE was designed as a speedometer, PhenoAge and GrimAge were designed as odometers, estimates of how much aging has occurred by the time of measurement^26^. We analyzed versions of the PhenoAge and GrimAge clocks calculated from DNA methylation principal components (“PC Clocks”), which have better technical reliability than to the original versions of these clocks^27^. PC Clocks were calculated using the R package available from GitHub (https://github.com/MorganLevineLab/PC-Clocks). We regressed clock-age values on participants’ chronological ages and computed residual values interpretable as how many more (or fewer) years of biological aging a person has experienced as compared the expectation based on their chronological age. For analysis, PhenoAge and GrimAge residuals were scaled to have *M*=0 and *SD*=1.

#### 2.2c Immune Cell Composition

Blood DNAm derives from white blood cells, with the precise mixture of different types varying between individuals. To account for the possibility that this heterogeneity could confound associations between epigenetic clocks and cognitive decline, we computed a set of control variables from the DNAm data to estimate the relative abundances of different cell types^28^. We computed values using the e*stimateCellCounts2* function from the *FlowSorted.BloodExtended.EPIC* R package developed by Salas et al. using the *preprocessNoob* setting on both this data and the cell reference dataset^29^. The package estimates relative abundances of 12 different types of immune cells (basophils, B naïve, B memory, CD4T naïve, CD4T memory, CD8T naïve, CD8T memory, eosinophils, monocytes, neutrophils, T regulatory cells and natural killer cells).

### 2.3 Neuropsychological Examination

All participants underwent annual standardized neuropsychological examinations beginning 19 to 47 years from study baseline and extending over 24 years of follow-up. We analyzed data from the Framingham Heart Study’s original neuropsychological battery^30^. The tests in this battery were organized into 8 cognitive domains according to factor analysis conducted by the Framingham Investigators: Verbal Memory (Logical Memory), Visual Memory (Visual Representation), Learning (Verbal Paired Associates)

Attention and Concentration (Trail Making Test), Abstract Reasoning (Similarities), Language (Boston Naming Test), Visuoperception (Hooper Visual Organization Test), and Psychomotor (Grooved Pegboard)^30^ (**Supplemental Table 1**). To integrate scores on these tests into a measure of global cognitive functioning, we followed the approach described by Downer et al^31^: First, we converted scores on each cognitive test to T-scores (*M*=50, *SD*=10) based on their distributions at baseline among individuals who remained free of dementia, Alzheimer’s Disease, and cerebrovascular disease through the end of follow-up (*N*=5010, 44% male, age: *M*=60 *SD*=16^32^). Next, we averaged the test-specific scores to compute our dependent variable for the main analysis-a measure of global cognitive functioning as defined by the Framingham Investigators^30,30^. Following established practices, global cognitive functioning scores were computed for participants with non-missing data on ≥70% of neuropsychological tests^33^.

#### 2.3a Cognitive Status Criteria

MCI was defined following the FHS Investigators’ practice as impaired performance by >1 *SD* on two or more cognitive tests in any domain^34,35^. Dementia status, subtype, and date of onset was defined by the FHS dementia review panel, which included serial assessments up to the time of death by staff neurologists and neuropsychologists, telephone interviews with caregivers, medical records, neuroimaging studies, and when available autopsies^32,36^. The diagnostic criteria are consistent with the Diagnostic and Statistical Manual of Mental Disorders, National Institute of Neurological and Communicative Disorders and Stroke and the Alzheimer’s Disease and Related Disorders Association, and The National Institute of Neurological Disorders and Stroke Association International pour la Recherche et l’Enseignement en Neurosciences.

#### 2.3b Cognitive Reserve

A prominent hypothesis in neuropsychology is that individuals vary in their cognitive resilience to neuropathology^37^. This phenomenon, referred to as cognitive reserve, could play a role as an effect-modifier in our analysis of pace of aging and cognitive decline. To explore this possibility, we tested baseline cognitive functioning as a modifier of associations between DunedinPACE and cognitive decline. For analysis, we computed average values across global cognition T-scores from neuropsychological assessments prior to DNAm baseline and dichotomized these average values at the healthy-population mean value of 50.

### 2.4 Smoking History

Smoking is known to affect blood DNA methylation and is also linked with cognitive decline^38,39^. To address potential confounding by smoking history, we created a composite index to summarize participants’ smoking history across the eight waves of follow-up prior to DNA methylation measurement. At each measurement wave, participants reported their smoking status as never, former, or current. We coded these responses as 0, 1, and 2, respectively, and averaged values across waves to form the final index.

### 2.6 APOE4

APOE4 status is a well-known risk factor for cognitive decline^40^. We assessed APOE4 as an effect modifier. APOE4 allele carrier status was coded dichotomously (one or more APOE4 allele versus no APOE4 allele).

### 2.6 Statistical Analysis

Analysis included *n*=2296 participants with DNA methylation data passing quality controls and measured global cognition at one or more assessments. We analyzed changes in global cognitive functioning using mixed-effects growth models implemented using the *lme4* package in the R software^41,42^. Our base model tested for change in cognition over time from DNAm and included covariates for age at baseline (linear and quadratic terms scaled in 10y units and centered at 65y), sex, a set of time-varying terms for follow-up time from DNAm (linear and quadratic terms scaled in 5y units) and the interaction of age at baseline with follow-up time (linear and quadratic).

#### 2.6a Testing pace of aging as a risk factor for cognitive decline

To test our hypothesis that faster pace of aging would predict more rapid cognitive decline, we added a term to the model for DunedinPACE and a product term modeling interaction between DunedinPACE and linear and quadratic follow-up time. The product terms tested association of DunedinPACE with rate of cognitive decline.

#### 2.6b Confounder adjustment

To address confounding by factors known to influence both cognitive decline and DNA methylation via pathways other than pace of aging, we repeated analysis adding covariates to the model for smoking history and leukocyte composition of blood samples used to derive DNA.

#### 2.6c Restriction of the analysis sample to participants who were cognitively intact at baseline

To evaluate sensitivity of results to patterns of decline among individuals already showing signs of impairment, we repeated analysis excluding individuals who manifested MCI prior to DNAm measurement .

#### 2.6d Effect modification

We conducted effect-modification analysis to evaluate contributions of cognitive-reserve processes, sex, and APOE4 carrier status to associations of pace of aging with cognitive decline. We tested effect modification by including main-effect terms for effect-modifiers along with product terms testing their interaction with follow-up time, DunedinPACE, and the Time*DunedinPACE term.

#### 2.6c Mediation

To test if more rapid cognitive decline mediated excess risk of dementia in individuals with faster pace of aging, we conducted formal mediation analysis using the survival analysis function within the CMAverse software (https://bs1125.github.io/CMAverse/).

#### 2.6d Other Clocks

We repeated analyses replacing DunedinPACE terms in our models with terms for the age-residuals of the PhenoAge and GrimAge epigenetic clocks.

## 3. RESULTS

We analyzed data for 2,296 non-Hispanic White adults in the Framingham Heart Study (FHS) Offspring Cohort followed for up to 23 years (*M* age=62, *SD*=9; 55% women; *M* global cognition T-score=51, *SD*=5, *Mdn* visits=4, *IQR* =2-12). At DNAm baseline, 23% of this sample met criteria for mild cognitive impairment; over follow-up, 12% were diagnosed with dementia, including Alzheimer’s disease. A comparison of the FHS Offspring sample analyzed here with the larger cohort is reported in **Table 1**.

**Table 1.** Characteristics of FHS Offspring Cohort and Analytic sample. The table reports characteristics of the Framingham Heart Study (FHS) Offspring Cohort and of the participants in that cohort included in our analysis. The analytic sample includes participants with DNA methylation data and complete data from at least one neuropsychological exam. Complete neuropsychological exam data were defined as >70 non-missing data across tests. Panel A reports demographic characteristics at study baseline (available for all participants). Panel B reports data on educational attainment (available for 77% of the Offspring Cohort and 99% of the Analytic Sample). Panel C reports data on incidence of dementia, Alzheimer’s disease, cerebrovascular disease, and mortality through the end of follow-up (available for 79% of the Offspring Cohort and 99% of the Analytic Sample).

|  | Offspring Cohort<br>*N* = 7306 | Analytic Sample<br>*n* = 2296 |
| --- | --- | --- |
|  | *M(SD) / %* |  |
| Chronological Age (years) | 61 (14) | 62 (9) |
| % Male | 44% | 45% |
| Education (years) | 15 (3) | 15 (2) |
| Global cognition (T-score) | 52 (5) | 51 (5) |
| % Dementia | 12% | 8% |
| % Alzheimer's disease | 9% | 5% |
| % Cerebrovascular disease | 2% | 2% |
| % Dead | 33% | 25% |

Participants’ global cognition scores declined over follow-up (per 5 years of follow-up, *B*=-1.54, 95%*CI*=[-1.66, -1.42]). Participants who were older at baseline experienced more rapid decline as compared to those who were younger; e.g., for those aged 65 at baseline, average 5y decline was -0.94 ([-1.06, -0.82], *p*<0.001); for those who were aged 75, it was much more rapid (age-by-time interaction *B*=-4.45 [-4.38, - 4.53], *p*<0.001).

### 3.1 Adults with faster pace of aging experienced more rapid cognitive decline

To test the hypothesis that participants who were experiencing a faster pace of biological aging would also exhibit more rapid cognitive decline, we tested associations of participants’ pace of aging with change over time in their global cognition scores.

Participants with a faster pace of aging tended to have worse average cognitive performance (DunedinPACE *B*=-0.92 [-1.16, -0.68], *p*<0.001) and more rapid cognitive decline over follow-up (DunedinPACE *B*=-0.21 [-0.32, -0.09], *p*<0.001).

Results were similar for the PhenoAge and GrimAge epigenetic clocks. Complete results for all clocks are shown in **Table 2**. Trajectories of cognitive aging for participants with slower and faster pace of aging/ older and younger biological age are illustrated in **Figure 1**.

**Table 2.** Epigenetic clock associations with cognitive functioning at baseline and cognitive decline over follow-up. The table shows results from mixed-effects regression analysis of changes in global cognition over up to 23 years of follow-up in the Framingham Heart Study Offspring Cohort (*n=*2296). Results are reported for three different epigenetic clocks (left column shows results for DunedinPACE; center column shows results for PhenoAge; right column shows results for GrimAge). Each set of results shows coefficient estimates, 95% CIs, and p-values for associations of epigenetic clocks with level of cognitive functioning at baseline (Intercept), linear slope of decline (Linear Slope), and quadratic slope of decline (Quadratic Slope). Results are reported for three sets of models. The base model included covariate adjustment for sex, age at baseline (quadratic), and associations of age at baseline with linear and quadratic slopes of cognitive decline. The second model added covariate adjustment for smoking history and DNA-estimated leukocyte composition. The third model excluded participants with mild cognitive impairment at DNA methylation baseline (*n*=528). Follow-up time was denominated in 5-year units. Coefficient estimates are denominated in global cognition T-score units per 1 SD of the epigenetic clocks.

|  | DunedinPACE |  |  | PhenoAge |  |  | GrimAge |  |  |
| --- | --- | --- | --- | --- | --- | --- | --- | --- | --- |
|  | *B* | *CI* | *p* | *B* | *CI* | *p* | *B* | *CI* | *p* |
| *Base Model* |  |  |  |  |  |  |  |  |  |
| Intercept | -0.92 | -1.16, -0.68 | **<0.001** | -0.66 | -0.89, -0.43 | **<0.001** | -0.92 | -1.16, -0.68 | **<0.001** |
| Linear Slope | -0.21 | -0.32, -0.09 | **<0.001** | -0.24 | -0.35, -0.12 | **<0.001** | -0.20 | -0.31, -0.09 | **<0.001** |
| Quadratic Slope | 0.03 | -0.03, 0.09 | 0.294 | 0.01 | -0.05, 0.07 | 0.782 | 0.01 | -0.05, 0.07 | 0.750 |
| *Adjusted for smoking history and DNAm-estimated leukocyte composition of DNA sample* |  |  |  |  |  |  |  |  |  |
| Intercept | -0.79 | -1.06, -0.53 | **<0.001** | -0.61 | -0.91, -0.31 | **<0.001** | -0.86 | -1.17, -0.54 | **<0.001** |
| Linear Slope | -0.21 | -0.32, -0.09 | **<0.001** | -0.24 | -0.35, -0.12 | **<0.001** | -0.20 | -0.31, -0.09 | **<0.001** |
| Quadratic Slope | 0.04 | -0.03, 0.10 | 0.258 | 0.01 | -0.05, 0.07 | 0.742 | 0.01 | -0.05, 0.07 | 0.725 |
| *Excluding participants with mild cognitive impairment at DNAm baseline* |  |  |  |  |  |  |  |  |  |
| Intercept | -0.76 | -0.99, -0.53 | **<0.001** | -0.58 | -0.81, -0.35 | **<0.001** | -0.79 | -1.02, -0.56 | **<0.001** |
| Linear Slope | -0.26 | -0.38, -0.13 | **<0.001** | -0.23 | -0.36, -0.11 | **<0.001** | -0.24 | -0.36, -0.12 | **<0.001** |
| Quadratic Slope | 0.05 | -0.01, 0.12 | 0.093 | 0.02 | -0.04, 0.08 | 0.533 | 0.02 | -0.04, 0.08 | 0.534 |

**Figure 1.**
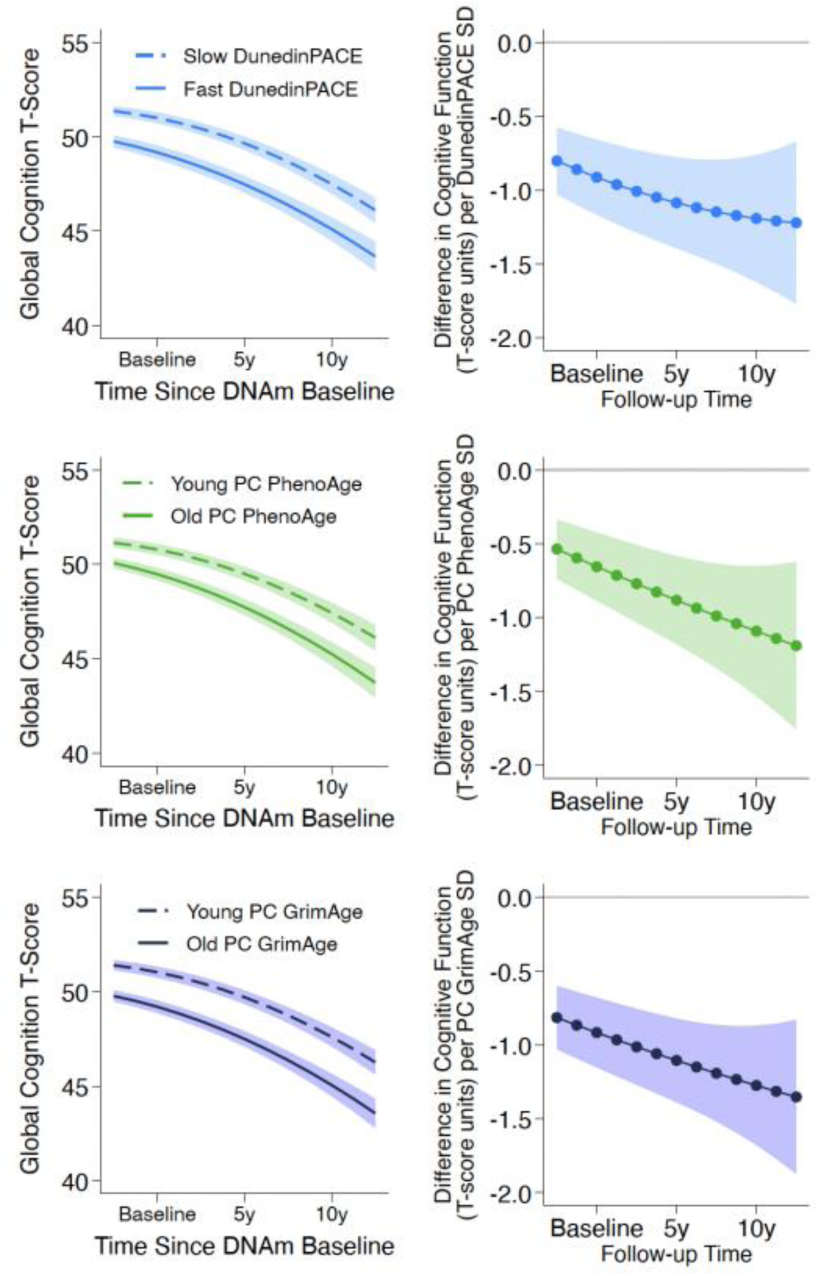
Predicted cognitive change over follow-up in Framingham Heart Study Offspring Cohort members with faster and slower pace of aging/ older and younger biological age. Data are plotted for analysis of three epigenetic clocks (*n*=2296). The top row shows analysis of the DunedinPACE epigenetic clock. The middle row shows analysis of the PhenoAge epigenetic clock. The bottom row shows analysis of the GrimAge epigenetic clock. Within each row, there are two graphs. The left-side graphs plot predicted trajectories of cognitive change over time for participants aged 65 at the time of DNAm collection. The Y axis shows cognitive functioning. The X axis shows follow-up time, centered at the time of DNAm collection. The dashed slope plots change for those with slow pace of aging/young biological age (one standard deviation below the mean). The solid slope plots change for those with fast pace of aging/old biological age (one standard deviation above the mean). The graphs show that participants with faster pace of aging/older biological age had poorer average cognitive functioning at baseline and experienced more rapid cognitive decline over follow-up as compared to those with slower pace of aging/younger biological age. The right-side graphs plot differences in cognitive functioning per epigenetic-clock SD over follow-up time. The graphs show that differences in cognitive functioning between those with faster/slower pace of aging and older/younger biological age increase with follow-up time.

We evaluated potential confounding of associations by smoking history and leukocyte composition of blood samples by adding covariates for these variables to our regression model. Results were similar to the primary model (**Table 2**).

Finally, we repeated analysis excluding individuals who manifested MCI at DNAm baseline. Results were similar to our primary model (**Tables 2**).

### 3.2 Exploration of effect modification by ADRD risk factors

We conducted exploratory analyses to evaluate sensitivity of associations between pace of aging and cognitive decline to modification by ADRD risk factors: cognitive reserve, sex, and APOE4 carrier status^37,40,43,44,45,46^. Cognitive reserve was not directly observed in our study. To conduct the sensitivity analysis, we grouped participants according to level of cognitive functioning at baseline (above/below a T-score of 50) to serve as a proxy of premorbid level of functioning. Participants with better baseline cognitive functioning were somewhat protected from risk associated with a faster pace of aging; being in the high-function group was associated with a reduction in the association of DunedinPACE with rate of cognitive decline by 36% (interaction b= 0.42, [0.19, 0.65], p<0.001). However, this effect modification was not observed for the PhenoAge and GrimAge clocks (interaction Bs<0.12, *p-values*>0.3). Trajectories of estimated marginal effects by year of follow-up for participants with slower and faster pace of aging and low and high cognitive reserve are graphed in **Figure 2**. Complete results for all clocks are reported in **Supplementary Table 2** and graphed in **Figure 2**. Associations of pace of aging with rate of cognitive decline were similar for men and women (interaction Bs<-0.12, *p-values*>0.3; **Supplemental Table 2**) and for carriers and non-carriers of APOE4 (interaction Bs<0.18, *p-values*>0.132; **Supplemental Table 2**).

**Figure 2.**
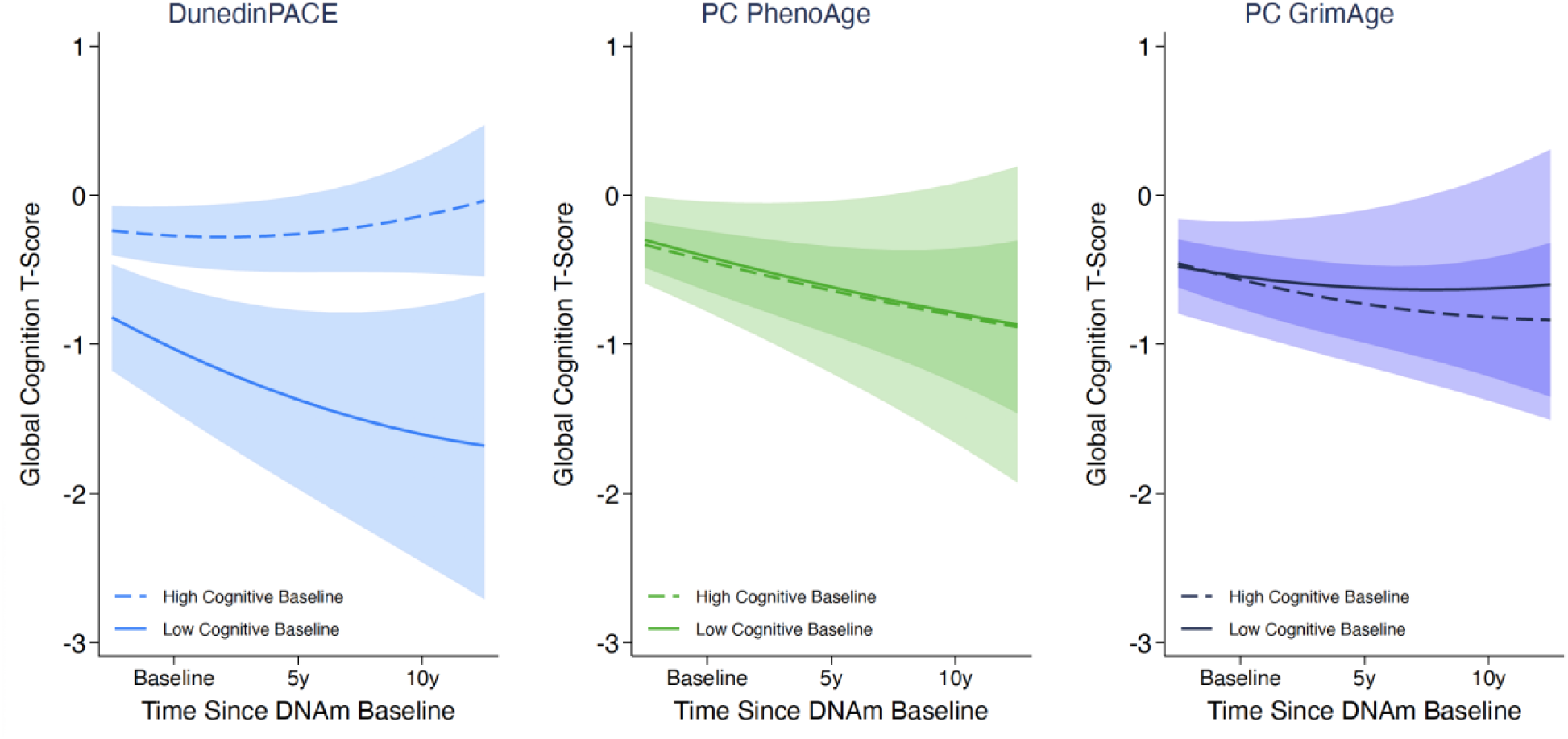
Associations of epigenetic clocks with cognitive change over follow-up in Framingham Heart Study Offspring Cohort members with high and low baseline cognitive functioning. The figure plots differences in cognitive functioning per epigenetic-clock SD over follow-up time for participants grouped by baseline cognitive functioning (*n*=2296). Data are plotted for analysis of three epigenetic clocks. The left graph shows analysis of the DunedinPACE epigenetic clock. The middle graph shows analysis of the PhenoAge epigenetic clock. The right graph shows analysis of the GrimAge epigenetic clock. Within each graph, the dashed slope shows the trend for participants with high baseline cognitive functioning (T≥50). The solid slope shows the trend for participants with low baseline cognitive functioning (T<50). DunedinPACE analysis shows that a faster pace of aging is associated with more rapid cognitive decline for those with low cognitive functioning at baseline, but this association is attenuated for participants with higher cognitive functioning at baseline. In contrast, in PhenoAge and GrimAge analysis, older biological age was associated with the same degree of decline regardless of baseline cognitive functioning.

### 3.3 Evaluation of dementia risk mediation

We previously reported that Framingham participants with faster pace of aging were at increased risk of developing dementia^47^. To integrate our current findings with this observation, we conducted mediation analysis. For mediation analysis, we restricted neuropsychological testing follow-up to the first three assessments following DNAm collection to avoid overlap with dementia diagnosis. Follow-up for mediation analysis included up to 14 years following DNAm collection. Over this period, 518 participants were diagnosed with dementia (mean follow-up to diagnosis= 9.34 years (SD=3.54).

Participants with faster DunedinPACE values had increased risk of dementia over follow-up (Total Effect HR=HR=1.62 [1.29, 2.03], p<0.001). Roughly 24% of this risk was mediated through accelerated cognitive decline over the first three assessments following DNAm collection (Indirect Effect HR=1.10 [1.05, 1.14], p<0.001). Full results are reported in **Supplemental Table 3**. Including covariate adjustment for level of cognitive functioning at baseline attenuated effect-sizes, but associations remained statistically different from zero (Total Effect HR=1.42 [1.21, 1.81], p=0.003; Indirect effect HR=1.05 [1.01, 1.09], p=0.011). Results were similar for PhenoAge and GrimAge, although associations of these clocks with dementia risk were smaller in magnitude (Total Effect HRs<1.21) and not statistically different from zero (p>0.06).

Complete results are reported in **Supplemental Table 3**.

A full comparison of clock effect-sizes across models is reported in **Supplemental Figure 2.**

## 4. DISCUSSION

We analyzed longitudinal neuropsychological testing data collected over two decades of follow-up in the Framingham Heart Study (FHS) Offspring Cohort to test if older adults with faster pace of biological aging experienced accelerated cognitive aging. We previously found that a faster pace of aging was associated with declines in IQ from childhood to midlife, signs of early brain aging, and earlier onset of dementia among older adults^14,15,25,47^. However, no data yet address whether faster pace of aging is associated with preclinical cognitive decline among older adults. In this study, we found that older adults with faster pace of aging as measured by the DunedinPACE epigenetic clock showed poorer cognitive functioning at baseline and experienced more rapid decline over follow-up. Our findings supported the robustness of this result across specifications considering a range of confounders and effect modifiers (e.g., analyses excluding participants with MCI). In mediation analysis, DunedinPACE associations with cognitive decline accounted for nearly a quarter of the overall relationship between DunedinPACE and dementia.

Our findings have implications for theory and research. With respect to theory, there are three implications. First, the extent to which cognitive decline reflects brain-specific or systemic processes is not fully understood. In previous studies, midlife and older adults with faster pace of aging exhibited brain characteristics linked with neuropathologies of aging, including cortical thinning and hippocampal atrophy^20,21^, suggesting connections between systemic aging and aging of the brain. This study complements those findings with evidence of corresponding decline in cognitive functioning. Together, our findings build the case that systemic biological aging contributes to the aging of the brain. The critical next steps are studies that can establish the temporal ordering of accelerated pace of aging, brain changes, and cognitive decline.

Second, whether biological aging contributes to dementia risk through an acceleration of preclinical cognitive decline versus increased risk of major neuropathologic events is unknown. We found that older adults with faster DunedinPACE experienced more rapid preclinical cognitive decline. Moreover, accelerated cognitive decline mediated roughly a quarter of the DunedinPACE association with dementia. Collectively, these results suggest a faster pace of biological aging contributes to accelerated preclinical cognitive decline and associated dementia risk. However, they also suggest that trajectories of preclinical decline are only one of multiple paths linking accelerated biological aging with dementia. Future studies should investigate the role of strokes and other neuropathological insults in linking faster pace of aging with increased dementia risk.

Third, sources of resilience to a faster pace of biological aging are unknown. In neuropsychology, the cognitive reserve hypothesis proposes that features of the brain that support cognitive functions buffer against cognitive decline in the face of accumulating neuropathology ^37,48^. A common approach to testing this hypothesis is to test effect-modification of cognitive decline by baseline levels of cognition. In FHS, we found that a faster pace of aging was less deleterious among older adults who had higher cognitive functioning at baseline as compared with adults who had poorer cognitive functioning at baseline. This result suggests pace of aging may relate to features of the brain promoting cognitive resilience to neuropathology. Studies are needed to identify the ways in which specific brain features may interact with pace of aging to affect trajectories of cognitive decline.

With respect to research, our findings contribute new evidence that an accelerated pace of aging is a harbinger of future dementia risk. The ADRD biomarker landscape is changing rapidly 50,51. If our findings can be replicated, DunedinPACE and other pace of aging measures may contribute to understanding the role systemic aging plays in ADRD pathogenesis. In clinical research, pace of aging measures could help identify individuals at risk for preclinical cognitive decline. Ultimately, tools like DunedinPACE could prove useful to clinicians treating cognitively intact older adults with subjective complaints, uncertain ADRD biomarker classifications, and ambiguous trajectories of cognitive decline. In the near term, DunedinPACE and related tools could enhance risk stratification for intervention studies ^49^. Finally, the connections between pace of aging and cognitive decline identified in our results suggests that interventions that slow pace of aging may also contribute to neuroprotection. As further evidence accumulates, DunedinPACE and related tools could provide near-term outcome measures for intervention studies seeking to modify life course accumulation of risk for ADRD.

We acknowledge limitations. There is no gold standard measure of biological aging.^1^ We focused on DunedinPACE based on three lines of evidence. First, DunedinPACE is predictive of diverse aging related outcomes, including disease, disability, and mortality^25^. Second, DunedinPACE is associated with social determinants of healthy aging in young, midlife, and older adults^25,52,53^. Third, DunedinPACE is modified by calorie restriction, an intervention that affects core biological processes of aging in animal experiments^54^. Generally, we saw similar effect sizes across DNAm epigenetic clocks, supporting the robustness of the findings. Our study relied on an observational design. Results do not establish causality of associations between DunedinPACE and cognitive decline. However, our longitudinal design does help establish temporal ordering of faster pace of aging and subsequent cognitive decline.

Our data do not establish which domains of cognitive functioning are most affected by pace of aging. The FHS neuropsychological battery includes only a single test in some domains and multiple tests in others; comparative analyses would be confounded by measurement artifacts^33^. The FHS Offspring Cohort we analyzed does not represent the US population. FHS recruited its participants in a single city in New England. The Offspring Cohort is overwhelmingly Non-Hispanic White. Moreover, it consists of participants whose families have been involved in biomedical research for multiple generations. Replication in more diverse cohorts, especially those representing populations at higher risk for ADRD, are essential to generalizing results from this study.

Trajectories of preclinical cognitive decline are well-established, but significant heterogeneity across individuals remains unexplained. Our study contributes evidence that an accelerated pace of biological aging is among the factors leading some individuals to experience more rapid trajectories of decline than others.

## Data Availability

All data produced in the present study are available upon reasonable request to the authors

## Supplemental Materials

### SUPPLEMENTAL METHODS

#### DNA Methylation Data

Prior to normalization, the following sample quality control was carried out. We removed n=38 samples with bisulfite conversion <80%, with methylated or unmethylated signal intensities <10.5 and mean detection p-values <0.005 or with <5 beads for >5% of probes, for which mean X- and Y-chromosome methylation levels of non-SNP CpGs were inconsistent with reported sex, with outlying values (<-4.0) for SNP-associated probes and thus a high probability of contamination or failure, that were clear outliers based on visual inspection of the first 2 principal components of autosome associated probes, and that failed any of the manufacturer’s thresholds for restoration, staining, extension, hybridization, target removal, bisulfite conversation of type I and II probes, specificity of type I and II probes, and intensity ratios among non-polymorphic probes (Fortin et al. 2017; Heiss and Just 2018). The remaining samples were normalized using the noob method (Fortin et al. 2017).

Fortin J-P, Triche TJ, Hansen KD. Preprocessing, normalization and integration of the Illumina HumanMethylationEPIC array with minfi. Bioinformatics. 2017; 33: 558–60.

Heiss JA, Just AC. Identifying mislabeled and contaminated DNA methylation microarray data: an extended quality control toolset with examples from GEO. Clin Epigenet. BioMed Central; 2018; 10: 1–9.

## SUPPLEMENTAL TABLES

**Supplemental Table 1.** Description of Neuropsychological Test Battery. This table describes the tests used to calculate global cognition across 8 cognitive domains, including Verbal Memory, Visual Memory, Learning, Attention and Concentration, Abstract Reasoning, Language, Visuoperception, and Psychomotor.

| Cognitive Domain | Test Measure | Description of Task |
| --- | --- | --- |
| Verbal Memory | Logical Memory – Immediate Recall | This task involves listening to and immediately recalling information from stories. The participants responses are scored for how accurately they recall details and themes of each story. |
|  | Logical Memory – Delayed Recall | 20-25 minutes following the immediate recall trials, the participant is asked to recall the stories again. |
|  | Logical Memory - % Retained | The percent of correct responses maintained across both the immediate and delayed recall trials. |
| Visual Memory | Visual Reproductions – Immediate Recall | This task involves observing and immediately drawing a replication of visual stimuli that includes various geometric shapes in a matrix. The participants responses are scored for how accurately they reproduce their shapes and their location on the page. |
|  | Visual Reproductions – Delayed Recall | 25 minutes following the immediate recall trial, the participant is asked to reproduce the visual stimuli again. |
|  | Visual Reproductions - % Retained | The percent of correct responses maintained across both the immediate and delayed recall trials. |
| Learning | Paired Associated - Total | A list of unrelated paired words are said aloud for the participant to learn. The participant is then prompted with one word and asked to recall its pair. The participant is scored on how accurately they can recall the paired word. |
| Attention and Concentration | Trail Making Test Part A – Time | Part A requires participants to connect a random arrangement of encircled numbers as quickly as possible. The participant is scored on how quickly and accurately they can complete the task. |
|  | Trail Making Test Part B - Time | Part B requires participants to repeat the task while alternating between numbers and letters (1 to A to 2 to B). The participant is scored on how |
|  |  | quickly and accurately they can complete the task. |
| Abstract Reasoning | Similarities – Total | This task involves giving the participant two words and asking how they are alike/similar. The participant is scored on how well they can reason the items likeness. |
| Language | Boston Naming Test – Total Correct Without cues | This task involves showing the participant a picture of an object and asking, “what is this?” The participant is scored on how accurately they can name the object without cueing. |
| Visuoperceptual Organization | Hooper Visual Organization Test – Total Score | This task involves showing the participant visual stimuli that has been broken up into puzzle-like pieces and asking them to identify what it is. The participant is scored on how accurately they can name the stimuli. |
| Psychomotor Speed | Finger Tapping – Dominant and Non-Dominant | The task involves asking the participant to tap their finger as quickly as possible on a dial for 10 seconds. The participant is scored on how many taps they can complete. |

**Supplemental Table 2.** Effect modification of epigenetic clock associations with cognitive functioning at baseline and cognitive decline over follow-up by sex, APOE4, and baseline cognition. The table shows results from mixed-effects regression analysis of changes in global cognition over up to 23 years of follow-up in the Framingham Heart Study Offspring Cohort (*n=*2296). Results are reported for three different epigenetic clocks (left column shows results for DunedinPACE; center column shows results for PhenoAge; right column shows results for GrimAge). For each epigenetic clock, results are reported for three sets of models. The first model tested effect modification by sex (male versus female). The second model tested effect modification by APOE4 (carrier versus non-carrier). The third model tested effect modification by baseline cognition (above or below T-score=50). For each model, coefficient estimates 95% CIs and p-values are reported for associations of epigenetic clocks, effect modifiers, and interaction terms with level of cognitive functioning at baseline (Intercept), linear slope of decline (Linear Slope), and quadratic slope of decline (Quadratic Slope, clocks only). All models included covariate adjustment for sex, age at baseline (quadratic), and associations of age at baseline with linear and quadratic slopes of cognitive decline. Follow-up time was denominated in 5-year units. Coefficient estimates are denominated in global cognition T-score units per 1 SD of the epigenetic clocks/level of the effect modifier.

|  | DunedinPACE |  |  | PhenoAge |  |  | GrimAge |  |  |
| --- | --- | --- | --- | --- | --- | --- | --- | --- | --- |
|  | B | CI | p | B | CI | p | B | CI | p |
| **Effect modification by sex** |  |  |  |  |  |  |  |  |  |
| *Clock Associations* |  |  |  |  |  |  |  |  |  |
| Intercept | -0.88 | -1.22, -0.55 | <0.001 | -0.65 | -1.00, -0.30 | <0.001 | -0.91 | -1.25, -0.57 | <0.001 |
| Linear Slope | -0.15 | -0.32, -0.02 | 0.077 | -0.22 | -0.40, -0.04 | 0.016 | -0.12 | -0.29, 0.05 | 0.169 |
| Quadratic Slope | 0.03 | -0.03, 0.10 | 0.279 | 0.01 | -0.05, 0.07 | 0.772 | 0.01 | -0.05, 0.07 | 0.716 |
| *Sex Associations* |  |  |  |  |  |  |  |  |  |
| Intercept | -0.32 | 0.76, 0.12 | 0.154 | -0.19 | -0.62, -0.30 | 0.399 | -0.67 | -1.14, -0.21 | 0.004 |
| Linear Slope | -0.04 | -0.48, 0.40 | 0.864 | 0.01 | -0.44, 0.45 | 0.982 | 0.00 | -0.47, 0.46 | 0.993 |
| *Sex\*Clock Associations* |  |  |  |  |  |  |  |  |  |
| Intercept | 0.13 | -0.08, 0.34 | 0.233 | 0.13 | -0.08, 0.34 | 0.219 | 0.06 | -0.17, 0.29 | 0.607 |
| Linear Slope | -0.08 | -0.30, 0.14 | 0.497 | -0.01 | -0.24, 0.22 | 0.933 | -0.12 | -0.36, 0.11 | 0.305 |
| **Effect modification by APOE4** |  |  |  |  |  |  |  |  |  |
| *Clock Associations* |  |  |  |  |  |  |  |  |  |
| Intercept | -0.92 | -1.16, -0.68 | <0.001 | -0.66 | -0.89, -0.43 | <0.001 | -0.92 | -1.16, -0.68 | <0.001 |
| Linear Slope | -0.21 | -0.32, -0.09 | <0.001 | -0.24 | -0.35, -0.12 | <0.001 | -0.20 | -0.31, -0.09 | <0.001 |
| Quadratic Slope | 0.03 | -0.03, 0.09 | 0.294 | 0.01 | -0.05, 0.07 | 0.782 | 0.01 | -0.05, 0.07 | 0.750 |
| *APOE4 Associations* |  |  |  |  |  |  |  |  |  |
| Intercept | -0.18 | -0.72, -0.32 | 0.504 | -0.20 | -0.74, 0.34 | 0.475 | -0.23 | -0.77, 0.31 | 0.410 |
| Linear Slope | 0.18 | -0.39, 0.75 | 0.539 | -0.07 | -0.61, 0.46 | 0.786 | 0.07 | -0.50, 0.65 | 0.805 |
| *APOE4\*Clock Associations* |  |  |  |  |  |  |  |  |  |
| Intercept | -0.56 | -0.83, -0.30 | <0.001 | -0.57 | -0.83, 0.30 | <0.001 | -0.57 | -0.84, -0.31 | <0.001 |
| Linear Slope | 0.18 | -0.11, 0.47 | 0.226 | -0.17 | -0.44, 0.10 | 0.208 | 0.00 | -0.29, 0.29 | 0.995 |
| **Effect modification by Baseline Cognition** |  |  |  |  |  |  |  |  |  |
| *Clock Associations* |  |  |  |  |  |  |  |  |  |
| Intercept | -1.07 | -1.35, -0.78 | <0.001 | -0.49 | -0.77, -0.21 | <0.001 | -0.60 | -0.87, -0.32 | <0.001 |
| Linear Slope | -0.46 | -0.65, -0.28 | <0.001 | -0.31 | -0.49, -0.13 | <0.001 | -0.19 | -0.36, -0.02 | 0.029 |
| Quadratic Slope | 0.04 | -0.02, 0.10 | 0.294 | 0.01 | -0.05, 0.07 | 0.691 | 0.02 | -0.03, 0.08 | 0.400 |
| *Baseline Cognition Associations* |  |  |  |  |  |  |  |  |  |
| Intercept | 7.06 | 6.66, 7.46 | <0.001 | 7.17 | 6.77, 7.57 | <0.001 | 7.12 | 6.72, 7.52 | <0.001 |
| Linear Slope | 0.24 | 0.00, 0.48 | 0.048 | 0.27 | 0.03, 0.51 | 0.027 | 0.26 | 0.02, 0.50 | 0.034 |
| *Baseline Cognition\*Clock Associations* |  |  |  |  |  |  |  |  |  |
| Intercept | 0.80 | 0.45, 1.15 | <0.001 | 0.06 | -0.29, 0.41 | 0.74 | 0.03 | -0.31, 0.38 | 0.852 |
| Linear Slope | 0.42 | 0.19, 0.65 | <0.001 | 0.12 | -0.11, 0.35 | 0.31 | 0.02 | -0.24, 0.21 | 0.883 |

**Supplemental Table 3.** Mediation analysis of epigenetic-clock associations with dementia risk. The table reports effect-size estimates from mediation models (*n=2296)*. Models evaluated mediation of epigenetic-clock associations with dementia risk by accelerated cognitive decline. Dementia risk was modeled using Cox time-to-event regression. Slope of cognitive decline was estimated over the first three neuropsychological assessments following DNA methylation baseline. Mediation analysis was conducted using CMAVerse R package. Results are reported for three different epigenetic clocks (left column shows results for DunedinPACE; center column shows results for PhenoAge; right column shows results for GrimAge). For each epigenetic clock, results are reported for two models. The first model included covariate adjustment for age at baseline, sex, and a product term modeling the interaction between age at baseline and follow-up time (linear and quadratic). The second model added a covariate for baseline cognitive functioning. Effect sizes are reported for total, direct, and indirect effects. Total effects reflect epigenetic clock associations with dementia. Direct effects reflect the portion of total effect that is independent of the mediator (slope of cognitive decline). Indirect effects reflect the portion of the total effect that is mediated by slope of cognitive decline. % mediated is calculated as the ratio of the indirect effect to the total effect.

|  | DunedinPACE |  |  | PhenoAge |  |  | GrimAge |  |  |
| --- | --- | --- | --- | --- | --- | --- | --- | --- | --- |
|  | HR | CI | p | HR | CI | p | HR | CI | p |
| *Base Model* |  |  |  |  |  |  |  |  |  |
| Total Effect | 1.69 | 1.29, 2.03 | <0.001 | 1.30 | 1.07, 1.57 | 0.007 | 1.36 | 1.09, 1.70 | <0.001 |
| Direct Effect | 1.47 | 1.81, 1.84 | <0.001 | 1.20 | 0.99, 1.44 | 0.056 | 1.24 | 1.00, 1.54 | 0.049 |
| Indirect Effect | 1.10 | 1.05, 1.15 | <0.001 | 1.08 | 1.04, 1.12 | <0.001 | 1.10 | 1.05, 1.41 | <0.001 |
| % Mediated | 23.5 | 11.8, 35.3 | <0.001 | 33.2 | 10.1, 56.3 | 0.005 | 32.7 | 10.8, 54.6 | 0.003 |
| *Adjusted for baseline cognition* |  |  |  |  |  |  |  |  |  |
| Total Effect | 1.42 | 1.21, 1.80 | 0.003 | 1.20 | 0.98, 1.47 | 0.066 | 1.18 | 0.93, 1.49 | 0.153 |
| Direct Effect | 1.35 | 1.07, 1.71 | 0.011 | 1.14 | 0.93, 1.39 | 0.195 | 1.13 | 0.89, 1.42 | 0.300 |
| Indirect Effect | 1.05 | 1.01, 1.09 | 0.010 | 1.05 | 1.02, 1.09 | 0.001 | 1.05 | 1.01, 1.08 | 0.008 |
| % Mediated | 16.1 | 2.3, 29.9 | 0.022 | 32.5 | -1.1, 66.0 | 0.058 | 30.2 | -10.6, 70.9 | 0.147 |

## Supplemental Figures

**Supplemental Figure 1.**
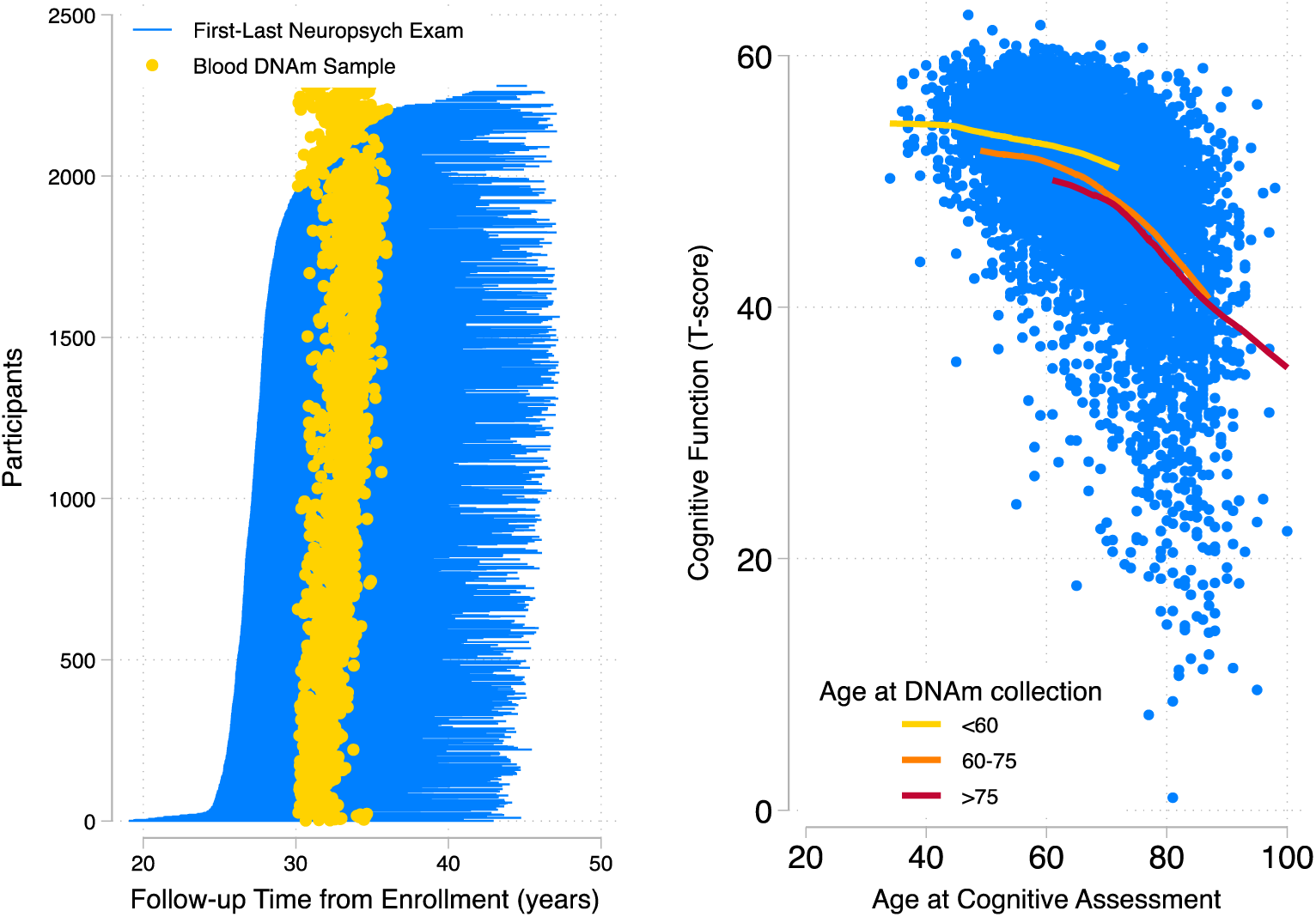
Cognitive decline with aging in the Framingham Heart Study Offspring Cohort. Panel A illustrates the timing of data collection. Each participant is represented as a blue line connecting their age at the time of their first neuropsychological exam to their age at their last neuropsychological exam. Participants are ordered according to the timing during follow-up of their first neuropsychological exam (from earliest at the bottom to latest at the top). Age at the time of the 8^th^ cohort follow-up visit, when blood DNA methylation (DNAm) data were collected, is marked with a gold circle. For most participants, neuropsychological examinations commenced around five years prior to the 8^th^ cohort follow-up visit. Panel B plots global cognitive functioning scores against chronological age at the time of testing. Each testing occasion is represented as an individual dot. The gold, orange, and red lines show locally-weighted regression slopes fitted to data for groups of participants defined by their age at the time of the 8^th^ study visit, when blood was collected for DNA methylation analysis.

**Supplemental Figure 2.**
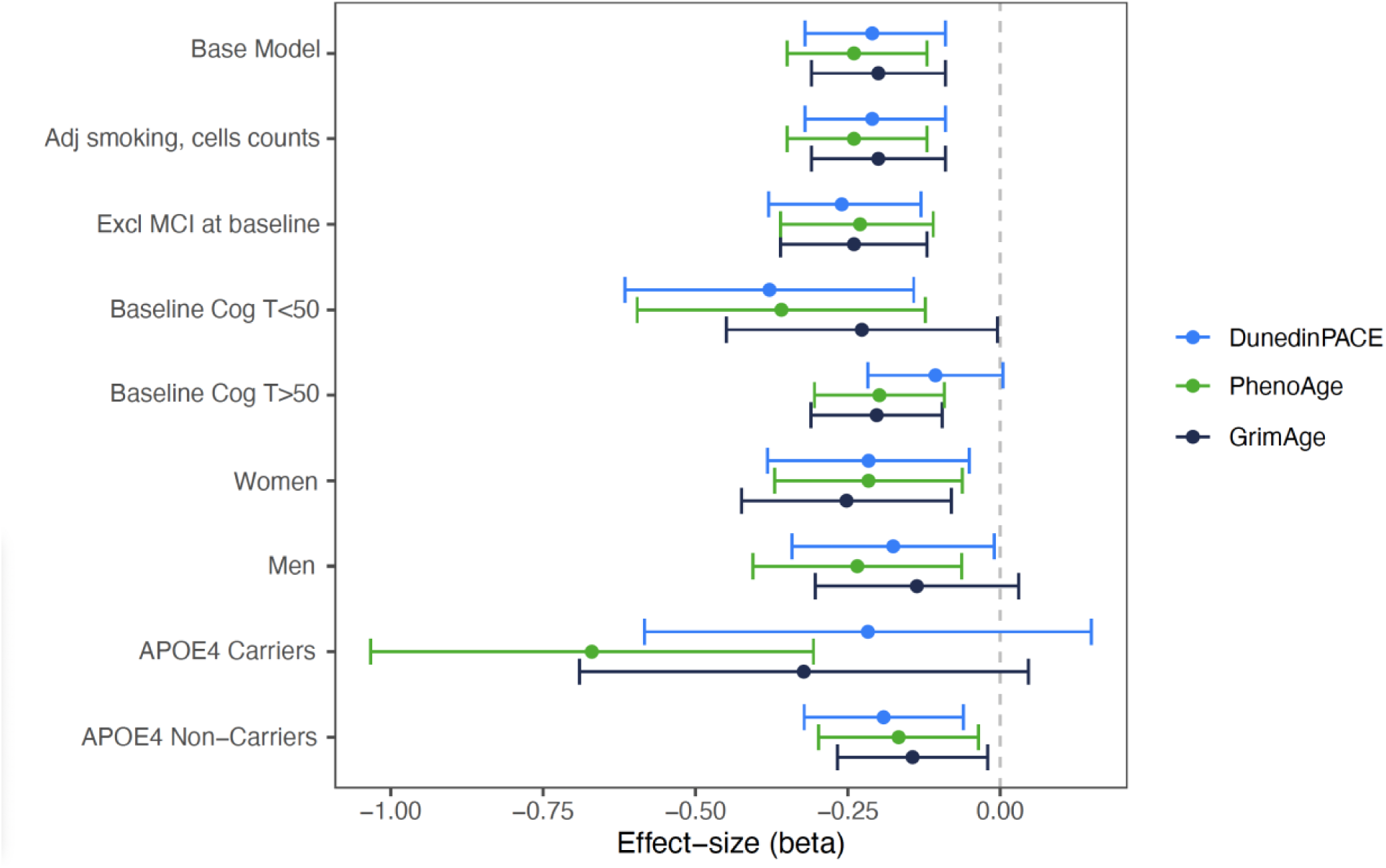
Effect sizes of predicted cognitive change over follow-up in Framingham Heart Study Offspring Cohort members across statistical models for each DNA methylation epigenetic clocks. This figure plots the effect sizes for predicted trajectories of cognitive change over time across various model specifications for each DNA methylation epigenetic clock (i.e., DunedinPACE, PhenoAge, GrimAge). The Y axis shows the various model specifications. The X axis shows follow-up time, centered at the time of DNAm collection. The blue dot graphs estimated change in cognition over five years for those with a faster DunedinPACE (one standard deviation above the mean). The green dot graphs change in cognition over five years for those with an older PhenoAge (one standard deviation above the mean). The black dot graphs change in cognition over five years for those with an older GrimAge (one standard deviation above the mean). This figure reports that we see similar effect sizes, or changes in cognition over time, across the various model specifications for each DNA methylation epigenetic clock.

